# Multidomain determinants of perivascular spaces indicate early cerebrovascular vulnerability in individuals at risk of Alzheimer’s disease

**DOI:** 10.64898/2026.03.23.26349050

**Authors:** Alba Fernandez-Bonet, Gerard Temprano-Sagrera, Patricia Genius, Blanca Rodriguez-Fernandez, Eugenia Domingo-Güell, Jordi Huguet, Mariateresa Buongiorno, Gonzalo Sanchez-Benavides, Marta Cirach, Mark Nieuwenhuijsen, Marleen de Bruijne, Tavia E. Evans, Natalia Vilor-Tejedor, the ALFA study

**Author notes:** **Corresponding Author: Gerard Temprano-Sagrera, PhD.** Barcelonaβeta Brain Research Center (BBRC), Pasqual Maragall Foundation. C. Wellington 30, 08005 Barcelona, Spain.; **Natalia Vilor-Tejedor, PhD.** Barcelonaβeta Brain Research Center (BBRC), Pasqual Maragall Foundation. C. Wellington 30, 08005 Barcelona, Spain. co-first authors (Alba Fernandez-Bonet, Gerard Temprano-Sagrera). The complete list of collaborators of the ALFA Study can be found in the acknowledgements section.

## Abstract

**Background:** Perivascular spaces (PVS) are increasingly recognized as MRI-visible markers of cerebrovascular and glymphatic functions. However, the determinants of regional PVS burden in the preclinical stages of Alzheimer’s disease (AD) remain to be elucidated, particularly when considering the impact of sex and core AD biomarkers.

**Methods:** We studied 1,199 cognitively unimpaired participants from the ALFA cohort, enriched for AD genetic risk and deeply phenotyped, including cerebrospinal fluid (CSF) biomarkers. PVS were automatically quantified in the basal ganglia, centrum semiovale, hippocampus, and midbrain on 3T MRI. Regional PVS counts were modeled using Poisson or negative binomial regression, accounting for demographics, cardiometabolic, lifestyle and genetic factors. Effect modification by sex and CSF amyloid/tau (AT) classification was examined.

**Results:** Determinants of PVS burden showed marked regional specificity. In the centrum semiovale, PVS burden was associated with age, male-sex and cardiometabolic factors. In contrast, hippocampal and midbrain PVS were more closely related to AD biomarker status and to psychological or sleep-related measures. Pronounced sex differences were observed, with men exhibiting higher PVS burden in centrum semiovale, while women showed higher PVS counts in the hippocampus, midbrain and basal ganglia. Associations between vascular, lifestyle, and genetic factors and PVS burden were stronger in amyloid-positive individuals, particularly those with concurrent amyloid and tau pathology, suggesting increased perivascular vulnerability along the AD continuum. Genetic risk scores for metabolic and lipid-related traits contributed modestly and independently to regional PVS variation, with evidence of sex- and biomarker-specific effects.

**Conclusions:** PVS burden reflects multidomain influences on cerebrovascular and clearance dysfunction in preclinical AD, with distinct regional, sex-specific, and biomarker-dependent patterns. These findings support PVS as an accessible MRI marker of early cerebrovascular vulnerability and highlight their potential relevance for risk stratification and precision prevention strategies before symptom onset.

## 1. Background

Perivascular spaces (PVS) are small, fluid-filled compartments that surround penetrating blood vessels in the brain and play a central role in interstitial fluid drainage, metabolic waste clearance, and neuroimmune regulation [1,2,3,4]. Once considered incidental findings, enlarged or magnetic resonance imaging (MRI)-visible PVS are now recognized as markers of cerebrovascular and glymphatic dysfunction and are increasingly studied in the context of brain aging and neurodegenerative diseases, including Alzheimer’s disease (AD) [5,6].

Quantitative MRI assessment of PVS provides a non-invasive window into cerebrovascular and glymphatic integrity [1,2,3]. Emerging evidence suggests that PVS burden exhibit marked regional heterogeneity across the brain, reflecting distinct pathophysiological mechanisms that may underlie PVS enlargement in different anatomical regions [7,8,9,10,11]. Increased PVS burden, particularly in the basal ganglia, may serve as an early imaging marker of vascular dysfunction and impaired brain clearance, processes increasingly linked to AD pathogenesis [1,12,13,14].

PVS burden is already considered a core neuroimaging feature of cerebral small vessel disease [15,16], and several studies have reported associations with cerebrovascular risk factors [17], Aβ and tau pathology [10,18], and neuroinflammatory mechanisms [2,19]. However, most existing evidence derives from general population samples or clinical cohorts with established cerebral small vessel disease or symptomatic neurodegeneration. Consequently, the determinants of PVS burden during the preclinical stages of AD, when cerebrovascular and clearance dysfunction may still be modifiable [7], remain poorly understood.

Current evidence suggests that PVS burden reflects the combined influence of demographic factors, vascular and metabolic health, lifestyle and environmental exposures, and genetic susceptibility [7,20,21]. Advanced age and male sex have consistently been associated with higher PVS burden, along with hypertension and obesity [1,7,17,22,23]. More recent evidence implicates environmental exposures [24], sleep disturbances [25,26] and neuropsychiatric symptoms [27,28] in glymphatic and cerebrovascular dysfunction. In parallel, genetic studies indicated that PVS burden is partly heritable and shares genetic architecture with cardiometabolic traits and AD-related pathways [21,29]. Yet, how these multidomain determinants shape regional PVS burden, and whether their effects differ by biological sex or AD biomarker status, remains largely unexplored.

Sex differences in vascular aging, lipid metabolism, hormonal regulation, sleep architecture, and neuroinflammatory responses may critically modify perivascular vulnerability, particularly in midlife. Likewise, the presence of early AD pathology, as defined by CSF Aβ and tau biomarkers, may alter the relationship between systemic risk factors and perivascular integrity. Understanding how sex and biomarker stage interact with multidomain risk factors is essential to disentangle early cerebrovascular and clearance dysfunction from downstream neurodegeneration and to identify tractable targets for prevention.

In this study, we examined demographic, cardiovascular, lifestyle/environmental and genetic determinants of PVS burden across four brain regions, in cognitively unimpaired participants from the ALFA study. We further investigated whether these associations varied by sex, CSF Aβ status and amyloid/tau (AT) biomarker classification. We hypothesized that PVS reflect the cumulative impact of genetic vulnerability and modifiable risk exposures, and that these effects are region- and stage-specific revealing early cerebrovascular vulnerability potentially influencing early AD pathology.

## 2. Methods

A schematic overview of the study design is presented in **Figure 1**.

**Fig 1.**
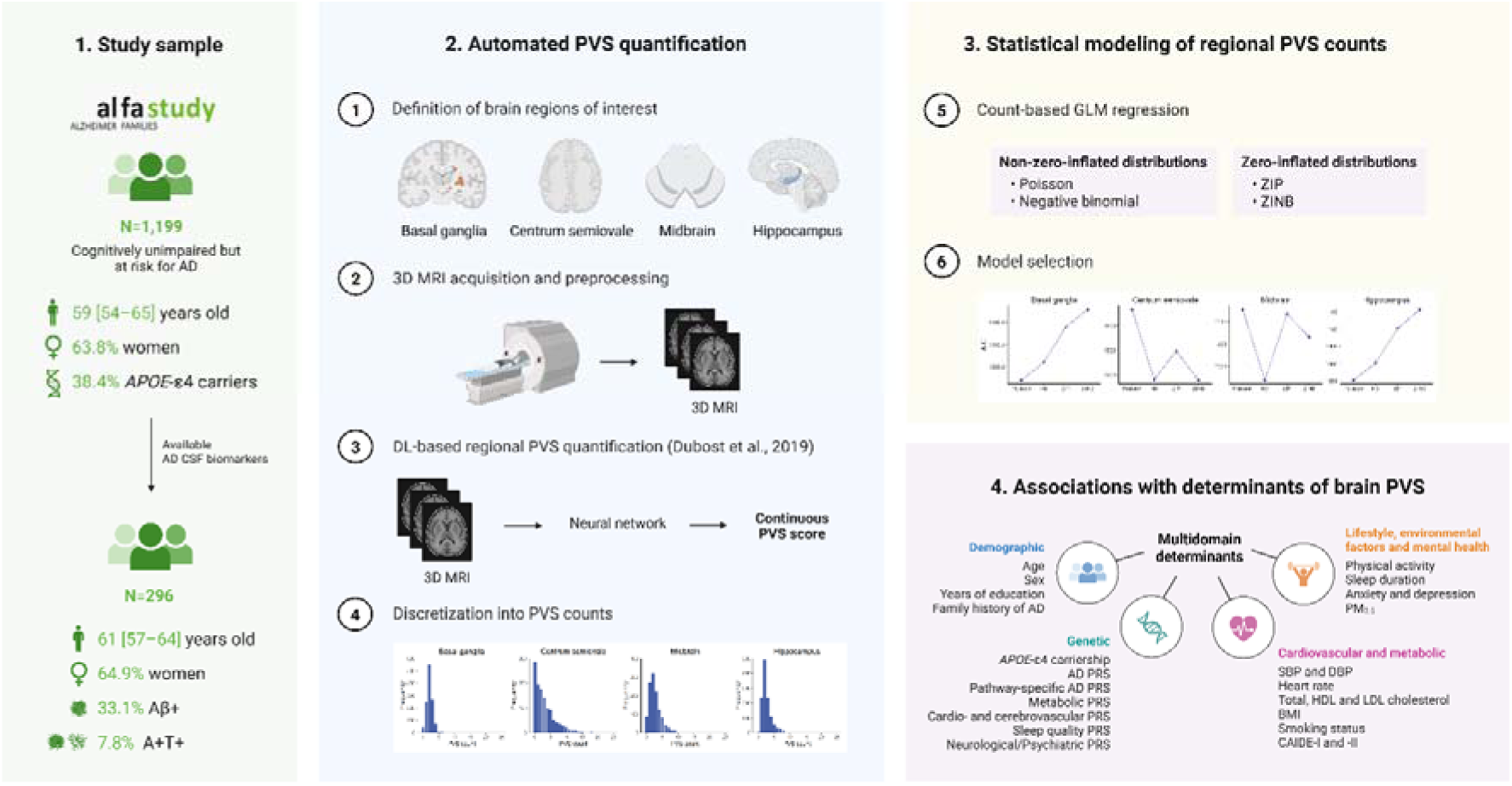
Study design overview. Schematic representation of the study workflow, including ALFA cohort sample description, MRI acquisition and automated PVS quantification, and multidomain determinant assessment. Analyses included main models in the whole sample, sex-stratified models, Aβ-stratified models, and AT biomarker–stratified models. All numerical estimates reported in the manuscript correspond to standardized rate ratios (RR) and RRs relative to the reference group for continuous and categorical determinants, respectively, with 95% confidence intervals (CI). Abbreviations: AD, Alzheimer’s Disease; BMI, Body Mass Index; CAIDE, Cardiovascular risk factors, Aging and Incidence of Dementia; CSF, Cerebrospinal Fluid; DBP, Diastolic Blood Pressure; DL, Deep Learning; GLM, Generalized Linear Model; HDL, High-Density Lipoprotein; LDL, Low-Density Lipoprotein; MRI, Magnetic Resonance Imaging; PM, Particulate Matter; PRS, Polygenic Risk Score; PVS, Perivascular Spaces; SBP, Systolic Blood Pressure; ZINB, Zero-Inflated Negative Binomial; ZIP, Zero-Inflated Poisson.

### Study Participants

Participants were drawn from the ALFA study, an AD genetically enriched research cohort designed to characterize early biological changes in the preclinical stage of AD [30]. The ALFA cohort includes CU adults, most of whom are first-degree descendants of patients with AD dementia, resulting in an elevated prevalence of *APOE*-ε4 carriers and higher polygenic susceptibility to AD [31]. For these analyses, we included 1,199 participants, aged 54-65 years, with available 3T structural MRI, genotyping, and comprehensive clinical and lifestyle assessments. Exclusion criteria were cognitive impairment, major neurological or psychiatric disorders, structural brain lesions, or poor MRI quality. A nested cohort of 296 participants had available CSF biomarker profiling that was used for biomarker-stratified analyses.

### MRI acquisition and Perivascular spaces quantification

Brain MRI data were acquired on a 3T scanner using harmonized ALFA imaging protocols [32]. The protocol included high-resolution T1- and T2-weighted sequences. Preprocessing steps included denoising, bias field correction, and spatial normalization. T1-weighted images were processed using the FreeSurfer (v6.0) recon-all pipeline to extract cortical and subcortical regions [33]. Regional brain volumes used for sensitivity analyses were derived from FreeSurfer (v7.1.1) [33] outputs: hippocampal and midbrain volumes were extracted directly from segmentation files, basal ganglia volume was computed as the sum of caudate, putamen, and pallidum, and centrum semiovale volume was approximated using total cerebral white matter volume.

PVS burden was automatically determined using a validated deep learning-based algorithm [34,35], and quantified in four bilateral regions of interest: basal ganglia, centrum semiovale, hippocampus, and midbrain. Regional PVS counts were averaged across hemispheres, and all model attention maps underwent quality-control to ensure anatomical accuracy. The automated quantification methodology has been described in detail by [35].

### Demographic, cardiovascular and lifestyle/environmental assessment

Demographic variables included age, sex, years of education, family history of AD and age of AD onset in first-degree relatives. Cardiovascular and metabolic risk factors were assessed through standardized clinical evaluations, including body mass index (BMI), systolic (SBP) and diastolic (DBP) blood pressure, resting heart rate, and serum lipid profiles (total cholesterol, high-density lipoprotein (HDL) cholesterol, and low-density lipoprotein (LDL) cholesterol). Cardiovascular dementia risk was quantified using the Cardiovascular risk factors, Aging and Incidence of Dementia (CAIDE) score [36], which integrates age, sex, education, BMI, SBP, DBP, total cholesterol, and physical activity (CAIDE-I), with an extended version that also incorporates *APOE*-ε4 carriership (CAIDE-II) [37]. Smoking status was assessed using standardized questionnaires (current, former or never), along with other lifestyle determinants information, physical activity (MET·hour/week), sleep duration (hour/day), and self-reported symptoms of anxiety and depression [38]. Exposure to particulate matter (PM_2.5_) air pollution was estimated with 2009 ESCAPE LUR models in Barcelona, based on seasonal measurements at 40 sites and geographic predictors [39]. Participants’ 2013–2014 addresses were geocoded to assign annual exposures, limited to stable residents (≥3 years) [40].

### Genetic data acquisition, quality control and polygenic risk scoring

Genomic DNA was extracted from peripheral blood samples and genotyped using the Illumina NeuroChip array [41]. Quality control followed established protocols [31], variants with call rate < 95%, minor allele frequency < 0.01, and Hardy-Weinberg equilibrium p-value < 10^-6^ were excluded. Samples with call rate < 98%, sex mismatch and excess heterozygosity were also excluded. Imputation was performed via the Michigan Imputation Server [42] using the Haplotype Reference Consortium Panel (HRC r1.1 2016). *APOE* genotypes were defined by rs429358 and rs7412 [43], and categorized as ε4 carriers and non carriers, as well as their ε4 genetic load (0/1/2 alleles). Polygenic risk scores (PRS) were computed in PRSice-2 using clumping and p-value thresholding [44], and were standardized to z-scores. PRS were derived for AD (AD, Aβ42, p-tau), cardio- and cerebrovascular diseases (hypertension, ischemic stroke, cerebral small vessel disease), metabolic traits (BMI, body fat index (BFI), HDL cholesterol, LDL cholesterol), sleep quality (insomnia, sleep duration), and neurological and psychiatric disorders (anxiety, major depressive, social interaction and isolation, stress), as well as pathway-specific mechanisms previously implicated in AD (amyloid, immune, external stimuli signaling, cholesterol efflux, complex lipoprotein metabolism) [45].

### Statistical Analysis

Regional PVS counts were inspected for distributional properties. Vuong’s non-nested test was applied to assess whether zero-inflated models provided a statistically superior fit over their standard counterparts. Model selection was further guided by log-likelihood values and Akaike Information Criterion (AIC). Based on these diagnostics, Poisson models were selected for basal ganglia and hippocampus regions, while negative binomial models were used for centrum semiovale and midbrain regions.

Each determinant domain (demographic; cardiovascular and metabolic; lifestyle, environmental factors and mental health; genetic) was analyzed separately, using generalized linear models adjusted for age, sex and intracranial volume.

Results were reported as rate ratios (RR) with 95% confidence intervals. False discovery rate (FDR) correction was applied within each domain to account for multiple comparisons. Effect modification by sex, Aβ status, and AT biomarker classification (A-T-, A+T-, A+T+) was examined through stratified models and interaction terms in the pooled sample to formally evaluate subgroup differences. Aβ positivity (A+) was defined as CSF Aβ42/40 < 0.071 [46]. Tau positivity (T+) was defined as CSF p-tau >24 mg/dL. A-T+ participants were excluded to filter out individuals who may not be within the AD *continuum*.

As a sensitivity analysis, all models were additionally adjusted for the volume of the corresponding anatomical region to account for potential confounding by regional brain size.

All analyses were performed in R version 4.3.1.

## 3. Results

### 3.1. Sample characteristics

The study sample included 1,199 CU participants (64% women) from the ALFA cohort, with a median age of 59.4 years. Sample characteristics are summarized in **Table 1**.

**Table 1.** Participant characteristics in the full sample and CSF biomarker subsample, stratified by sex, Aβ status, and AT biomarker group. Variables include demographic, cardiovascular and metabolic, lifestyle and genetic factors, and regional PVS counts. Continuous variables are shown as median (IQR) and categorical variables are presented as n (%). P-values were obtained using the Kruskal-Wallis test for continuous variables, and the χ² test or Fisher’s exact test for categorical variables, as appropriate. Sample size varies across variables due to missing data. The CSF biomarker subsample includes participants with available CSF amyloid and tau measurements. Legend: N, Sample Size; AD, Alzheimer’s disease; CSF, cerebrospinal fluid; PM2.5 Particulate Matter; SBP, Systolic Blood Pressure; DBP, Diastolic Blood Pressure; HDL, High-Density Lipoprotein Cholesterol; LDL, Low-Density Lipoprotein Cholesterol; BMI, Body Mass Index; CAIDE, Cardiovascular risk factors, Aging and Incidence of Dementia; PVS, Perivascular Spaces.

|  | Sex |  |  |  |  | Aβ status |  |  |  |  | AT classification |  |  |  |  |  |
| --- | --- | --- | --- | --- | --- | --- | --- | --- | --- | --- | --- | --- | --- | --- | --- | --- |
|  | N | Total | Men (N=434) | Women (N=765) | P-value | N | Total | A- (N=198) | A+ (N=98) | P-value | N | Total | A-T- (N=198) | A+T- (N=75) | A+T+ (N=23) | P-value |
| Demographic determinants |  |  |  |  |  |  |  |  |  |  |  |  |  |  |  |  |
| Age (years), median (IQR) | 1199 | 59.41<br>(53.79–64.61) | 59.72<br>(53.74–64.91) | 59.26<br>(53.81–64.51) | 0.526 | 296 | 60.61<br>(56.71–64.07) | 59.94<br>(56.27–63.71) | 62.47<br>(58.11–65.54) | 0.003 | 296 | 60.61<br>(56.71–64.07) | 59.94<br>(56.27–63.71) | 61.12<br>(57.72–64.94) | 64.78<br>(62.41–66.95) | <0.001 |
| Sex, n (%) | 1199 |  |  |  | - | 296 |  |  |  | 0.593 | 296 |  |  |  |  | 0.563 |
| Male |  | 434 (36.2) | - | - |  |  | 104 (35.1) | 67 (33.8) | 37 (37.8) |  |  | 104 (35.1) | 67 (33.8) | 30 (40.0) | 7 (30.4) |  |
| Female |  | 765 (63.8) | - | - |  |  | 192 (64.9) | 131 (66.2) | 61 (62.2) |  |  | 192 (64.9) | 131 (66.2) | 45 (60.0) | 16 (69.6) |  |
| Education (years), median (IQR) | 1199 | 12.00<br>(11.00–17.00) | 15.00<br>(12.00–17.00) | 12.00<br>(10.00–17.00) | <0.001 | 296 | 12.00<br>(11.00–17.00) | 12.00<br>(11.00–17.00) | 12.00<br>(11.00–17.00) | 0.708 | 296 | 12.00<br>(11.00–17.00) | 12.00<br>(11.00–17.00) | 15.00<br>(11.00–17.00) | 11.00<br>(8.50–15.00) | 0.051 |
| Family history of AD, n (%) | 1199 |  |  |  | 0.456 | 296 |  |  |  | 0.581 | 296 |  |  |  |  | 0.866 |
| No |  | 128 (10.7) | 42 (9.7) | 86 (11.2) |  |  | 20 (6.8) | 15 (7.6) | 5 (5.1) |  |  | 20 (6.8) | 15 (7.6) | 4 (5.3) | 1 (4.3) |  |
| Yes |  | 1071 (89.3) | 392 (90.3) | 679 (88.8) |  |  | 276 (93.2) | 183 (92.4) | 93 (94.9) |  |  | 276 (93.2) | 183 (92.4) | 71 (94.7) | 22 (95.7) |  |
| Age of onset in family history of AD (years), median (IQR) | 1136 | 72.00<br>(68.00–76.00) | 71.00<br>(67.00–76.00) | 72.00<br>(68.00–76.00) | 0.152 | 288 | 72.00<br>(67.00–76.00) | 71.00<br>(67.00–76.00) | 73.00<br>(66.75–75.00) | 0.859 | 288 | 72.00<br>(67.00–76.00) | 71.00<br>(67.00–76.00) | 73.00<br>(69.00–75.00) | 70.00<br>(65.50–75.00) | 0.578 |
| Cardiovascular and metabolic risk determinants |  |  |  |  |  |  |  |  |  |  |  |  |  |  |  |  |
| SBP (mmHg), median (IQR) | 939 | 126.00<br>(114.50–138.50) | 130.50<br>(122.00–141.50) | 122.50<br>(111.50–136.00) | <0.001 | 73 | 129.00<br>(121.00–142.50) | 128.50<br>(119.75–140.62) | 129.00<br>(122.00–145.50) | 0.283 | 73 | 129.00<br>(121.00–142.50) | 128.50<br>(119.75–140.62) | 130.00<br>(122.38–147.75) | 125.00<br>(118.50–140.25) | 0.352 |
| DBP (mmHg), median (IQR) | 939 | 79.00<br>(72.25–86.00) | 82.00<br>(76.50–87.75) | 77.00<br>(70.00–84.00) | <0.001 | 73 | 79.00<br>(72.00–86.50) | 78.75<br>(71.38–82.62) | 80.50<br>(72.00–88.50) | 0.433 | 73 | 79.00<br>(72.00–86.50) | 78.75<br>(71.38–82.62) | 81.75<br>(72.12–89.62) | 74.00<br>(69.50–83.50) | 0.473 |
| Heart rate (bpm), median (IQR) | 938 | 68.50<br>(62.00–76.00) | 66.00<br>(59.50–74.50) | 70.00<br>(64.00–76.50) | <0.001 | 73 | 65.50<br>(60.00–75.00) | 67.75<br>(62.75–75.75) | 65.00<br>(59.00–73.50) | 0.271 | 73 | 65.50<br>(60.00–75.00) | 67.75<br>(62.75–75.75) | 65.25<br>(59.12–73.25) | 65.00<br>(59.00–75.00) | 0.545 |
| Total cholesterol (mg/dL), median (IQR) | 321 | 204.00<br>(183.00–228.00) | 191.00<br>(175.00–210.00) | 209.50<br>(190.00–232.75) | <0.001 | 296 | 204.00<br>(183.00–228.25) | 204.00<br>(185.00–225.75) | 203.50<br>(180.00–229.00) | 0.574 | 296 | 204.00<br>(183.00–228.25) | 204.00<br>(185.00–225.75) | 203.00<br>(180.50–229.00) | 204.00<br>(177.50–222.50) | 0.685 |
| HDL cholesterol (mg/dL), median (IQR) | 316 | 61.50<br>(52.00–73.00) | 53.00<br>(45.00–62.00) | 66.00<br>(57.00–77.00) | <0.001 | 291 | 62.00<br>(52.00–73.00) | 62.00<br>(51.00–73.25) | 61.00<br>(53.00–71.50) | 0.672 | 291 | 62.00<br>(52.00–73.00) | 62.00<br>(51.00–73.25) | 60.50<br>(53.00–71.00) | 66.00<br>(54.50–72.50) | 0.608 |
| LDL cholesterol (mg/dL), median (IQR) | 304 | 131.00<br>(112.00–151.25) | 125.00<br>(109.75–144.25) | 136.00<br>(115.00–152.00) | 0.004 | 279 | 131.00<br>(112.00–151.50) | 132.50<br>(112.75–151.25) | 128.00<br>(111.50–151.00) | 0.763 | 279 | 131.00<br>(112.00–151.50) | 132.50<br>(112.75–151.25) | 128.00<br>(109.00–155.50) | 125.50<br>(116.75–145.50) | 0.933 |
| BMI (kg/m²), median (IQR) | 1199 | 26.03<br>(23.81–28.80) | 26.90<br>(24.87–29.23) | 25.43<br>(23.14–28.25) | <0.001 | 296 | 26.03<br>(23.73–28.79) | 25.90<br>(23.68–28.73) | 26.05<br>(23.82–28.80) | 0.813 | 296 | 26.03<br>(23.73–28.79) | 25.90<br>(23.68–28.73) | 26.02<br>(23.85–28.79) | 27.59<br>(23.47–29.15) | 0.888 |
| Smoking status, n (%) | 1050 |  |  |  | 0.140 | 266 |  |  |  | 0.511 | 266 |  |  |  |  | 0.498 |
| Never smoked |  | 188 (17.9) | 61 (15.8) | 127 (19.1) |  |  | 51 (19.2) | 30 (17.2) | 21 (22.8) |  |  | 51 (19.2) | 30 (17.2) | 15 (20.8) | 6 (30.0) |  |
| Current smoker |  | 276 (26.3) | 94 (24.4) | 182 (27.4) |  |  | 61 (22.9) | 42 (24.1) | 19 (20.7) |  |  | 61 (22.9) | 42 (24.1) | 17 (23.6) | 2 (10.0) |  |
| Former smoker |  | 586 (55.8) | 230 (59.7) | 356 (53.5) |  |  | 154 (57.9) | 102 (58.6) | 52 (56.5) |  |  | 154 (57.9) | 102 (58.6) | 40 (55.6) | 12 (60.0) |  |
| CAIDE-I (0–15), median (IQR) | 1052 | 5.00<br>(4.00–7.00) | 6.00<br>(5.00–8.00) | 5.00<br>(4.00–6.00) | <0.001 | 293 | 5.00<br>(4.00–6.00) | 5.00<br>(4.00–6.00) | 6.00<br>(4.00–7.00) | 0.616 | 293 | 5.00<br>(4.00–6.00) | 5.00<br>(4.00–6.00) | 5.00<br>(4.00–6.75) | 6.00<br>(4.50–6.50) | 0.668 |
| CAIDE-II (0–15), median (IQR) | 1030 | 7.00<br>(5.00–9.00) | 8.00<br>(6.00–9.00) | 7.00<br>(5.00–8.00) | <0.001 | 293 | 8.00<br>(6.00–9.00) | 7.00<br>(5.75–9.00) | 8.00<br>(7.00–10.00) | <0.001 | 293 | 8.00<br>(6.00–9.00) | 7.00<br>(5.75–9.00) | 8.00<br>(7.00–10.00) | 8.00<br>(7.00–9.50) | 0.002 |
| Lifestyle, environmental factors and mental health determinants |  |  |  |  |  |  |  |  |  |  |  |  |  |  |  |  |
| Physical activity (MET·h/week), median (IQR) | 1038 | 33.20<br>(15.62–58.42) | 38.50<br>(19.13–68.34) | 29.33<br>(14.62–52.42) | <0.001 | 262 | 33.43<br>(16.20–58.22) | 33.50<br>(16.26–58.00) | 32.75<br>(16.46–58.22) | 0.750 | 262 | 33.43<br>(16.20–58.22) | 33.50<br>(16.26–58.00) | 36.15<br>(20.86–59.21) | 19.52<br>(9.43–43.69) | 0.084 |
| Sleep duration (h/day), median (IQR) | 1054 | 7.00<br>(6.50–8.00) | 7.00<br>(6.30–7.00) | 7.00<br>(7.00–8.00) | 0.024 | 268 | 7.00<br>(7.00–7.50) | 7.00<br>(6.50–7.50) | 7.00<br>(7.00–7.62) | 0.446 | 268 | 7.00<br>(7.00–7.50) | 7.00<br>(6.50–7.50) | 7.00<br>(7.00–7.00) | 7.00<br>(7.00–8.00) | 0.134 |
| Anxiety, n (%) | 1198 |  |  |  | 0.290 | 296 |  |  |  | 0.958 | 296 |  |  |  |  | 0.883 |
| No |  | 1080 (90.2) | 397 (91.5) | 683 (89.4) |  |  | 273 (92.2) | 182 (91.9) | 91 (92.9) |  |  | 273 (92.2) | 182 (91.9) | 70 (93.3) | 21 (91.3) |  |
| Yes |  | 118 (9.8) | 37 (8.5) | 81 (10.6) |  |  | 23 (7.8) | 16 (8.1) | 7 (7.1) |  |  | 23 (7.8) | 16 (8.1) | 5 (6.7) | 2 (8.7) |  |
| Depression, n (%) | 1198 |  |  |  | 0.001 | 296 |  |  |  | 0.730 | 296 |  |  |  |  | 0.430 |
| No |  | 1084 (90.5) | 409 (94.2) | 675 (88.4) |  |  | 271 (91.6) | 180 (90.9) | 91 (92.9) |  |  | 271 (91.6) | 180 (90.9) | 71 (94.7) | 20 (87.0) |  |
| Yes |  | 114 (9.5) | 25 (5.8) | 89 (11.6) |  |  | 25 (8.4) | 18 (9.1) | 7 (7.1) |  |  | 25 (8.4) | 18 (9.1) | 4 (5.3) | 3 (13.0) |  |
| PM<sub>2.5</sub> (µg/m³), median (IQR) | 975 | 14.98<br>(14.12–16.68) | 15.05<br>(14.26–16.92) | 14.95<br>(14.06–16.51) | 0.071 | 236 | 15.07<br>(14.16–16.89) | 15.17<br>(14.23–16.86) | 14.99<br>(14.16–16.94) | 0.817 | 236 | 15.07<br>(14.16–16.89) | 15.17<br>(14.23–16.86) | 14.99<br>(14.13–16.60) | 15.04<br>(14.50–19.00) | 0.611 |
| Genetic determinants |  |  |  |  |  |  |  |  |  |  |  |  |  |  |  |  |
| APOE-ε4 carriership, n (%) | 1181 |  |  |  | 0.712 | 294 |  |  |  | <0.001 | 294 |  |  |  |  | <0.001 |
| No |  | 727 (61.6) | 260 (60.7) | 467 (62.0) |  |  | 145 (49.3) | 123 (62.8) | 22 (22.4) |  |  | 145 (49.3) | 123 (62.8) | 13 (17.3) | 9 (39.1) |  |
| Yes |  | 454 (38.4) | 168 (39.3) | 286 (38.0) |  |  | 149 (50.7) | 73 (37.2) | 76 (77.6) |  |  | 149 (50.7) | 73 (37.2) | 62 (82.7) | 14 (60.9) |  |
| APOE-ε4 load, n (%) | 1163 |  |  |  | 0.709 | 290 |  |  |  | <0.001 | 290 |  |  |  |  | <0.001 |
| Non-carrier |  | 713 (61.3) | 256 (60.2) | 457 (61.9) |  |  | 142 (49.0) | 120 (62.5) | 22 (22.4) |  |  | 142 (49.0) | 120 (62.5) | 13 (17.3) | 9 (39.1) |  |
| One ε4 allele |  | 394 (33.9) | 150 (35.3) | 244 (33.1) |  |  | 122 (42.1) | 62 (32.3) | 60 (61.2) |  |  | 122 (42.1) | 62 (32.3) | 47 (62.7) | 13 (56.5) |  |
| Two ε4 alleles |  | 56 (4.8) | 19 (4.5) | 37 (5.0) |  |  | 26 (9.0) | 10 (5.2) | 16 (16.3) |  |  | 26 (9.0) | 10 (5.2) | 15 (20.0) | 1 (4.3) |  |
| **Perivascular spaces** |  |  |  |  |  |  |  |  |  |  |  |  |  |  |  |  |
| Basal ganglia PVS, median (IQR) | 1199 | 2.00<br>(2.00–3.00) | 2.00<br>(1.00–2.00) | 2.00<br>(2.00–3.00) | **<0.001** | 296 | 2.00<br>(1.75–3.00) | 2.00<br>(1.00–3.00) | 2.00<br>(2.00–3.00) | **0.023** | 296 | 2.00<br>(1.75–3.00) | 2.00<br>(1.00–3.00) | 2.00<br>(1.00–3.00) | 2.00<br>(2.00–3.00) | 0.052 |
| Centrum semiovale PVS, median (IQR) | 1199 | 2.00<br>(1.00–4.50) | 3.00<br>(2.00–6.00) | 2.00<br>(0.00–3.00) | **<0.001** | 296 | 2.00<br>(0.00–4.25) | 2.00<br>(0.00–4.00) | 2.00<br>(1.00–5.00) | 0.091 | 296 | 2.00<br>(0.00–4.25) | 2.00<br>(0.00–4.00) | 2.00<br>(1.00–5.00) | 4.00<br>(2.00–6.00) | **0.042** |
| Midbrain PVS, median (IQR) | 1199 | 2.00<br>(1.00–3.00) | 2.00<br>(1.00–3.00) | 2.00<br>(1.00–4.00) | **<0.001** | 296 | 2.00<br>(1.00–4.00) | 2.00<br>(1.00–4.00) | 2.00<br>(2.00–4.00) | 0.499 | 296 | 2.00<br>(1.00–4.00) | 2.00<br>(1.00–4.00) | 2.00<br>(2.00–4.00) | 2.00<br>(1.50–4.00) | 0.774 |
| Hippocampus PVS, median (IQR) | 1199 | 2.00<br>(2.00–3.00) | 2.00<br>(1.00–2.00) | 2.00<br>(2.00–4.00) | **<0.001** | 296 | 2.00<br>(2.00–3.00) | 2.00<br>(2.00–3.00) | 3.00<br>(2.00–3.75) | **0.018** | 296 | 2.00<br>(2.00–3.00) | 2.00<br>(2.00–3.00) | 3.00<br>(2.00–3.00) | 2.00<br>(2.00–4.00) | 0.059 |

### 3.2 Main analysis

Detailed estimates for all models are shown in **Figure 2** and **Supplementary Table 1**.

**Fig 2.**
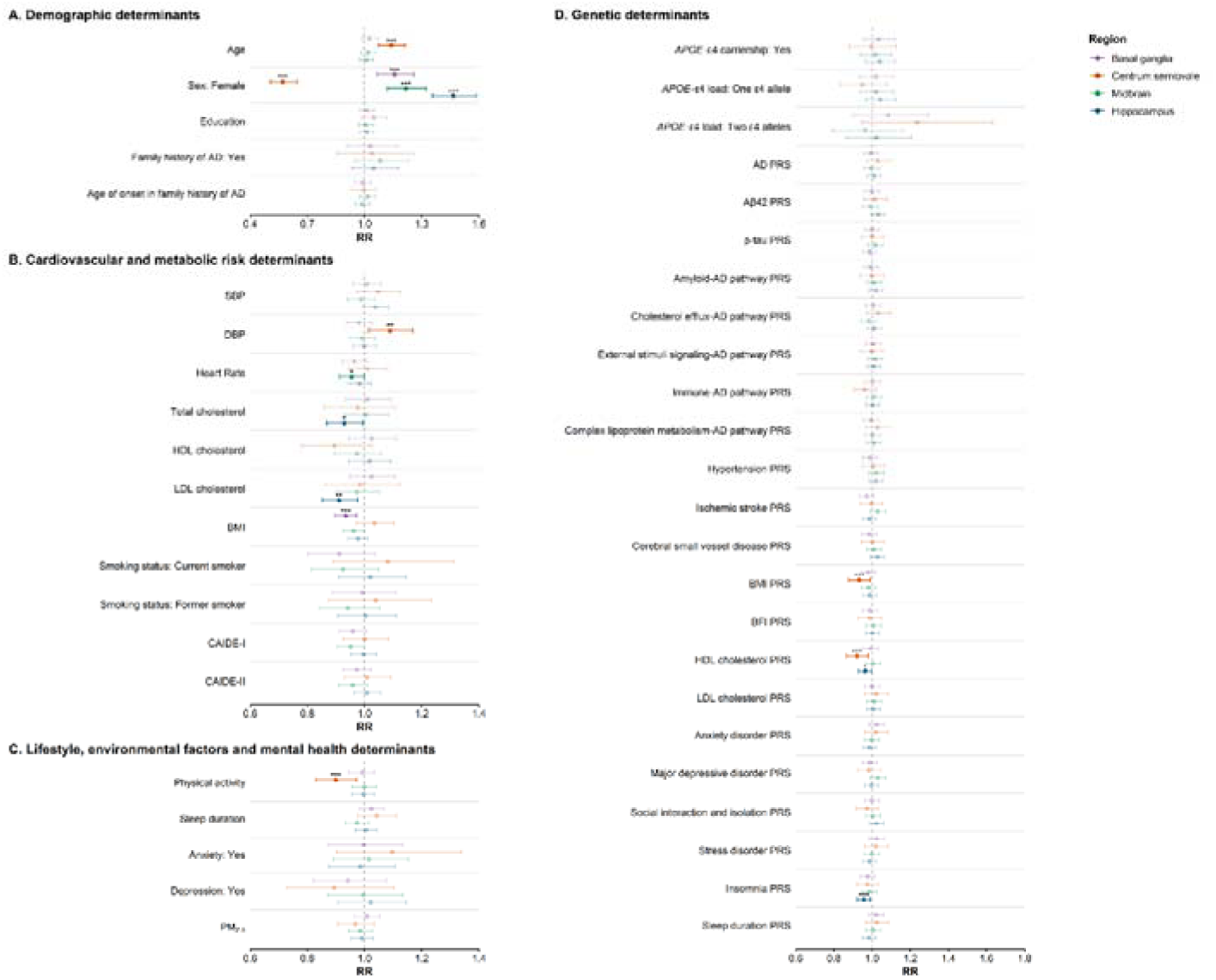
Main models. Associations of demographic, cardiovascular and metabolic, lifestyle, and genetic determinants with regional PVS counts in the full sample (N=1,199). Each panel represents one brain region (basal ganglia, centrum semiovale, midbrain, hippocampus). Models were fitted using Poisson or negative binomial regression, as appropriate. Results are displayed as standardized rate ratios (RR) and as RRs relative to the reference group for continuous and categorical variables, respectively, with 95% CIs. Significance: *** pFDR<0.05; ** p<0.025; * p<0.05.

#### Demographic determinants

Higher PVS counts were significantly associated with older age and male sex, consistent with previous literature. Each additional year of age corresponded to a ∼1-2% increase in centrum semiovale PVS counts (95% CI=1.073–1.213, *p*_FDR_*=*3.48×10^-5^), independent of sex. Men exhibited higher PVS counts than women in the centrum semiovale (95% CI=0.505–0.644, *p*_FDR_*=*5.07×10^-19^), whereas women showed higher PVS counts in the other regions (95% CI=1.356–1.586, *p*_FDR_*=*5.13×10^-21^ in the hippocampus; 95% CI=1.120–1.323, *p*_FDR_*=*1.77×10^-5^ in the midbrain; 95% CI=1.065–1.257, *p*_FDR_*=*0.002 in the basal ganglia). Years of education, as a proxy for cognitive reserve, was not significantly associated with PVS counts in any region.

#### Cardiovascular and metabolic risk determinants

Higher DBP was nominally associated with increased PVS counts at the centrum semiovale (95% CI=1.015–1.169, *p=*0.015), while elevated LDL cholesterol levels (95% CI=0.851–0.975, *p*=0.007) and total cholesterol levels (95% CI=0.867–0.995, *p*=0.037) were associated with lower hippocampal-PVS burden. In contrast, higher BMI was associated with reduced basal ganglia PVS counts (95% CI=0.897–0.973, *p*_FDR_=0.012).

#### Lifestyle, environmental factors and mental health determinants

Higher physical activity was associated with reduced centrum semiovale PVS counts (95% CI=0.831–0.971, *p*_FDR_*=*0.033). Other lifestyle factors and psychological variables, including smoking, sleep duration, and anxiety or depression showed nominal trends or region-specific trends but did not survive multiple comparison corrections.

#### Genetic determinants

PRS for BMI and PRS for HDL cholesterol were significantly associated with lower centrum semiovale PVS counts (95% CI=0.878–0.990, *p*_FDR_*=*0.043; 95% CI=0.866–0.979, *p*_FDR_*=*0.030, respectively), while the HDL cholesterol PRS was associated with lower hippocampal PVS counts (95% CI=0.930–0.999, *p*=0.045). A PRS for insomnia was also associated with lower hippocampus-PVS counts (95% CI=0.923–0.991, *p*_FDR_*=*0.030).

### 3.3 Sex-specific effect modification analyses

Following the pronounced sex-based disparities observed in PVS counts across all regions, we conducted sex-interactions and stratified analyses to identify specific differences between women and men [**Figure 3**, **Supplementary Figure 1**].

**Fig 3.**
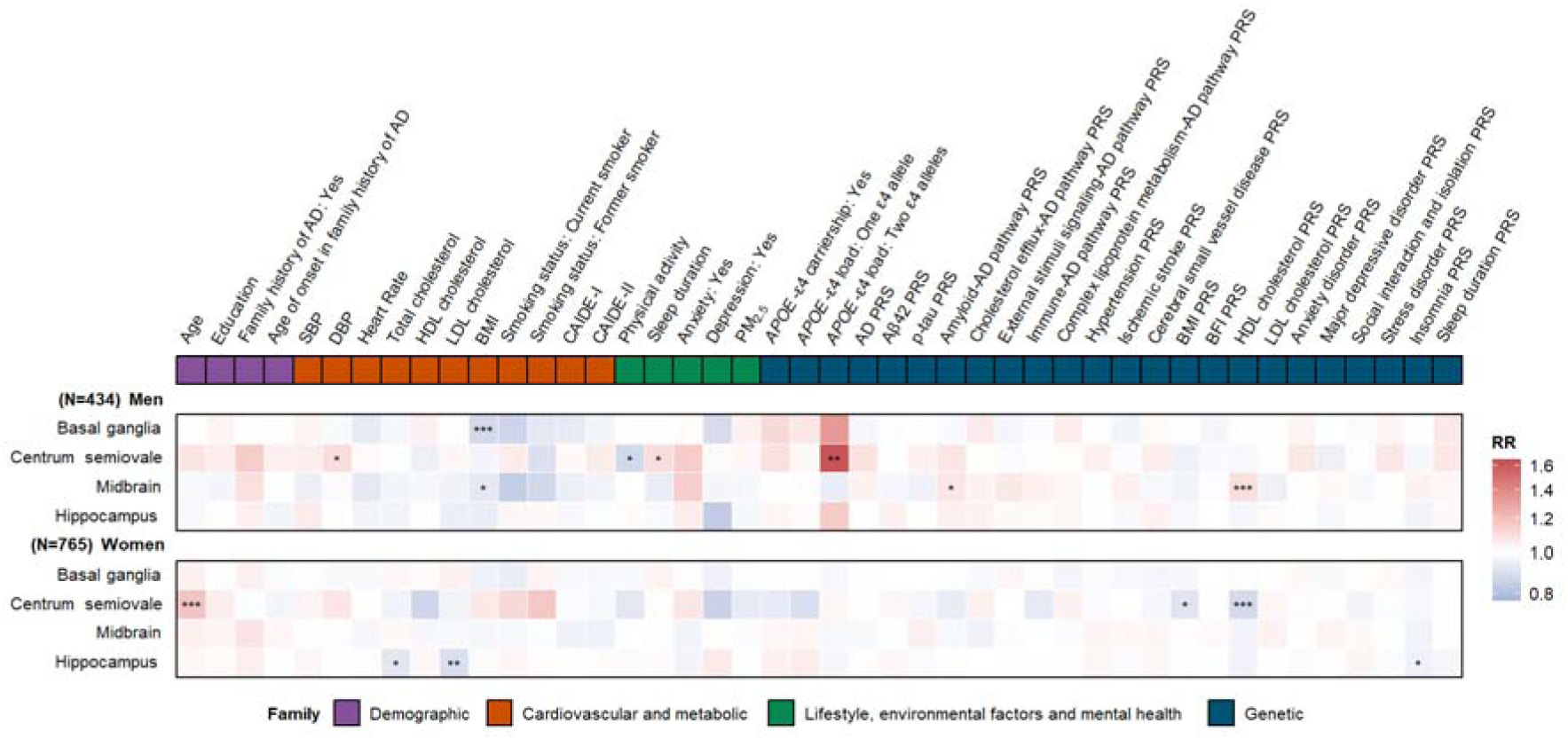
Sex-stratified models. Associations between multidomain determinants and regional PVS counts stratified by sex. Panel A: Women. Panel B: Men. Models follow the same analytical framework as in Figure 2 and were fitted independently in each sex group. Estimates and 95% CIs were obtained from Poisson or negative binomial models, as appropriate, and represent standardized rate ratios (RR) and RRs relative to the reference group for continuous and categorical variables, respectively. Significant sex interactions corresponding to these results are shown in Supplementary Table 2. Significance: *** pFDR<0.05; ** p<0.025; * p<0.05.

A significant interaction with the PRS for HDL cholesterol was observed in the midbrain (*p*_FDR_=0.015), suggesting sex-specific lipid-related genetic influences on perivascular integrity [**Supplementary Table 2**]. Nominal interactions were also found for BMI and PRS for complex lipoprotein metabolism pathway in the basal ganglia (*p*=0.025, *p*=0.041, respectively), PRS for p-tau in the midbrain (*p*=0.037), and PRS for amyloid pathway in both the centrum semiovale (*p*=0.027) and midbrain (*p*=0.025).

Sex-stratified analyses showed that, in men, higher PRS for HDL cholesterol was significantly associated with greater midbrain PVS counts (95% CI=1.025–1.170, *p*_FDR_=0.027), consistent with the interaction effect noted above. Higher BMI was significantly associated with lower PVS counts in the basal ganglia (95% CI=0.817–0.946, *p*_FDR_=0.007) [**Supplementary Table 3**].

In women, PVS counts were not significantly associated with any of the determinants that showed significant sex-interaction terms. However, other significant and nominal associations were observed, predominantly in the centrum semiovale PVS count. Age showed a positive association with greater PVS burden (95% CI=1.085–1.292, *p*_FDR_=3.20×10^-4^). A higher PRS for HDL cholesterol was associated with reduced PVS counts (95% CI=0.809–0.964, *p*_FDR_=0.017) [**Supplementary Table 4**].

### 3.4 Aβ-specific effect modification analyses

Psychological and behavioral factors showed significant interactions with Aβ status, including sleep duration (*p*_FDR_=0.044) and depressive symptoms (*p*_FDR_=0.022) in the hippocampus [**Figure 4**, **Supplementary Figure 2**, **Supplementary Table 5**]. Anxiety also showed a nominal interaction in the centrum semiovale (*p*=0.025). Other nominal interactions were observed between Aβ status and several vascular and lifestyle determinants. Specifically, interactions were found for SBP (hippocampus, *p*=0.027) and PM_2.5_ air pollution exposure (basal ganglia, *p*=0.048). Within the genetic domain, a nominal interaction was found for cholesterol efflux AD pathway PRS (*p*=0.047) in the midbrain.

**Fig 4.**
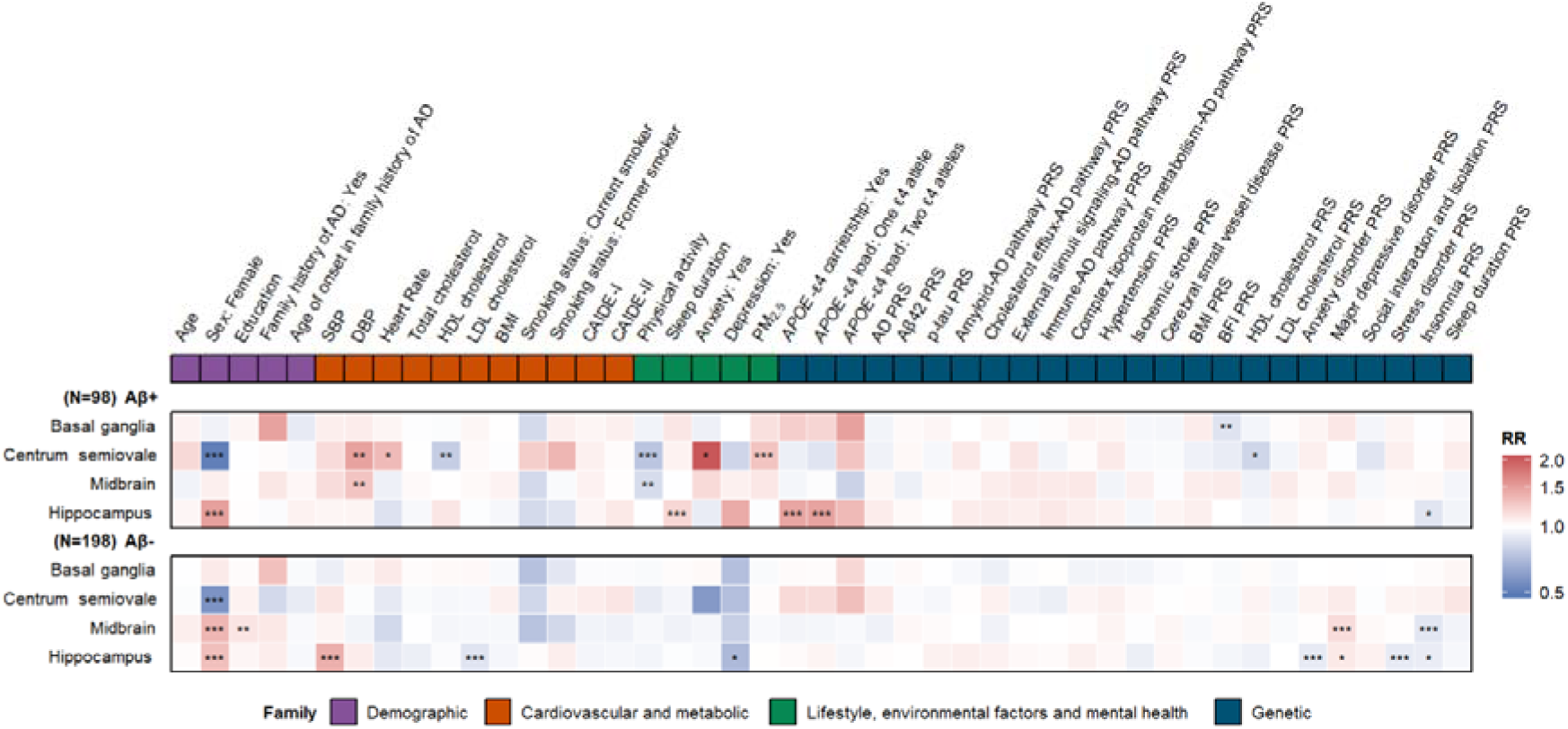
Aβ-stratified models. Associations between multidomain determinants and regional PVS counts stratified by CSF amyloid status. Panel A: Aβ- individuals. Panel B: Aβ+ individuals. Models follow the same analytical framework as in Figure 2 and were fitted independently in each Aβ group. Estimates and 95% CIs were obtained from Poisson or negative binomial models, as appropriate, and represent standardized rate ratios (RR) and RRs relative to the reference group for continuous and categorical variables, respectively. Significant determinant–Aβ interactions are summarized in Supplementary Table 5. Significance: *** pFDR<0.05; ** p<0.025; * p<0.05.

Aβ-stratified analyses showed that, in Aβ+ individuals, female sex remained a robust determinant of PVS burden, presenting a significantly lower centrum semiovale-PVS counts (95% CI=0.343–0.749, *p*_FDR_=0.003) and higher hippocampal PVS counts (95% CI=1.171–1.958, *p*_FDR_=0.009). Among lifestyle factors, physical activity was associated with reduced centrum semiovale-PVS burden (95% CI=0.599–0.917, *p*_FDR_=0.031), whereas longer sleep duration was associated with higher hippocampal PVS counts (95% CI=1.051–1.345, *p*_FDR_=0.031). Increased PM_2.5_ exposure (95% CI=1.057–1.583, *p*_FDR_=0.042) was also linked to greater centrum semiovale-PVS burden. In the genetic domain, *APOE*-ε4 carriership and ε4 heterozygosity were significantly associated with greater hippocampal PVS burden (95% CI=1.086–2.018, *p*_FDR_=0.046; 95% CI=1.097–2.050, *p*_FDR_=0.046, respectively) **[Supplementary Table 6].**

In Aβ- individuals, sex-related differences were consistent, with female sex being associated with lower centrum semiovale and higher hippocampal and midbrain PVS counts (95% CI=0.410–0.783, *p*_FDR_=0.003; 95% CI=1.089–1.602, *p*_FDR_=0.026; 95% CI=1.104–1.699, *p*_FDR_=0.022, respectively). Within the vascular domain, increased SBP was associated with higher hippocampal PVS counts (95% CI=1.106–1.811, *p*_FDR_=0.033), while LDL cholesterol was associated with lower PVS counts in the hippocampus (95% CI=0.803–0.955, *p*_FDR_=0.028). Distinct genetic associations emerged in Aβ- individuals. PRS for anxiety and stress were negatively associated with hippocampal PVS counts (95% CI=0.824–0.985, *p*_FDR_=0.044 for both), while PRS for major depressive disorder was positively associated with midbrain PVS counts (95% CI=1.047–1.285, *p*_FDR_=0.019). PRS for insomnia showed a negative association with midbrain PVS counts (95% CI = 0.797–0.972, *p*_FDR_=0.022) [**Supplementary Table 7**].

### 3.5 AT-specific effect modification analyses

Nominal interactions were observed between CSF-determined AT biomarker classification and several vascular, lifestyle, and psychological determinants. These findings suggest that the influence of systemic and behavioral factors on PVS burden may vary across AD biomarker stages [**Figure 5**, **Supplementary Figure 3**, **Supplementary Table 8**]. Nominally significant interactions (A+T- vs. A-T-) were found for heart rate in the centrum semiovale (*p*=0.044), SBP in the hippocampus (*p*=0.035), ambient PM_2.5_ exposure in the basal ganglia (*p*=0.034), and anxiety in the centrum semiovale (*p*=0.010). In addition, significant interactions emerged (A+T+ vs. A-T-) for sleep duration (*p*=0.012) and depressive symptoms (*p*=0.006) in the hippocampus, and the cholesterol efflux AD pathway PRS (*p*=0.04) in the midbrain. Smoking status also showed nominal interactions with both former smoking in the centrum semiovale (*p*=0.010) and current smoking in the midbrain (*p*=0.011).

**Fig 5.**
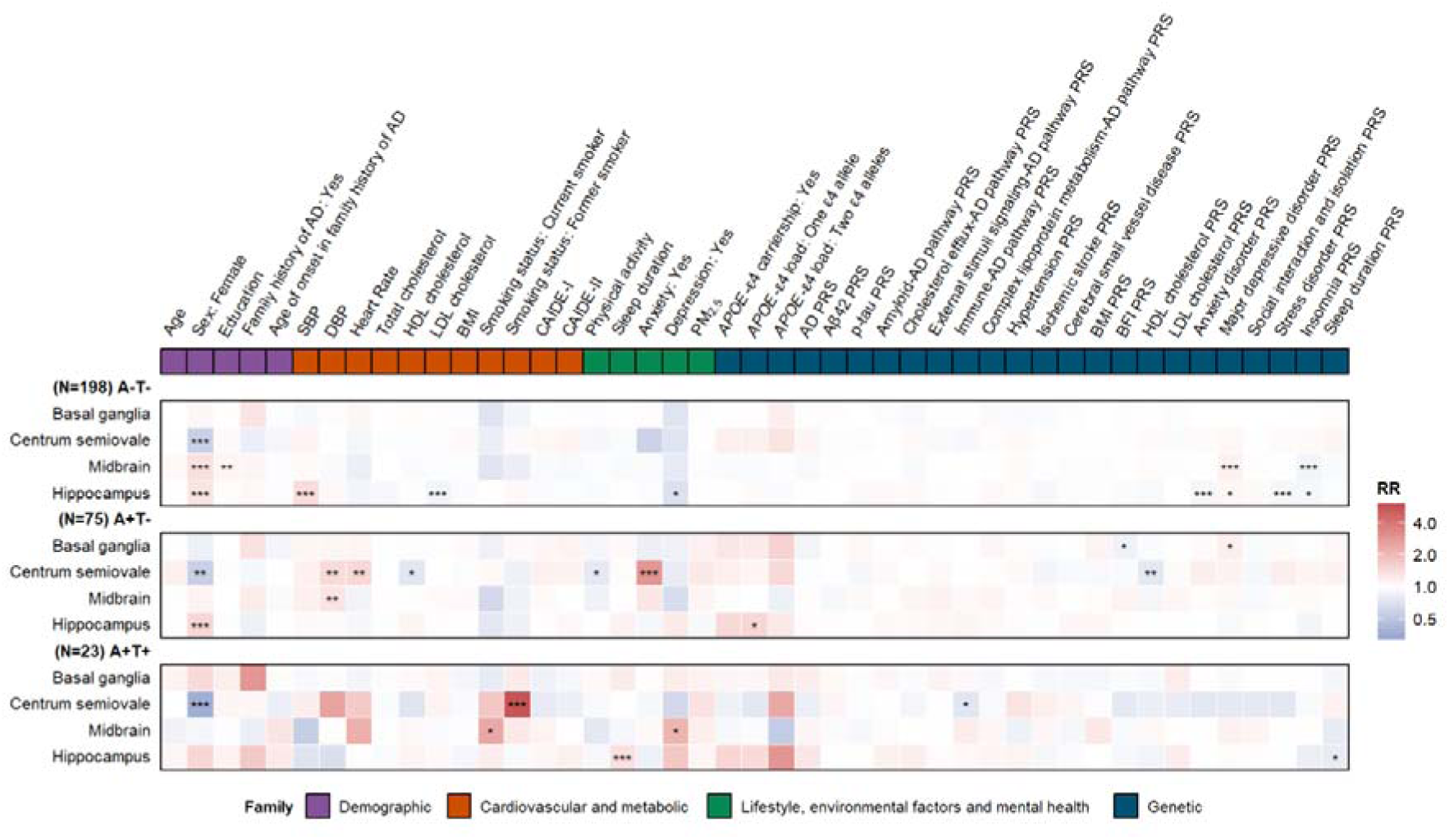
AT biomarker group–stratified models. Associations between multidomain determinants and regional PVS counts across AT biomarker stages. Panel A: A-T- group. Panel B: A+T- group. Panel C: A+T+ group. Models follow the same analytical framework as in Figure 2 and were fitted independently in each AT biomarker group. Estimates and 95% CIs were obtained from Poisson or negative binomial models, as appropriate, and represent standardized rate ratios (RR) and RRs relative to the reference group for continuous and categorical variables, respectively. For clarity, upper CI bounds outside the plotting range were truncated. Interaction effects between determinants and AT groups are reported in Supplementary Table 8. Significance: *** pFDR<0.05; ** p<0.025; * p<0.05.

AT-stratified analyses showed that, in participants without evidence of amyloid or tau pathology (A-T-), female sex remained a consistent determinant, and was associated with lower centrum semiovale PVS counts (95% CI=0.410–0.783, *p*_FDR_=0.003), and higher PVS counts in the hippocampus (95% CI=1.089–1.602, *p*_FDR_= 0.026) and midbrain (95% CI=1.104–1.699, *p*_FDR_=0.022). Within the cardiovascular and metabolic domain, increased SBP was associated with higher hippocampal PVS counts (95% CI=1.106–1.811, *p*_FDR_=0.033), whereas higher LDL cholesterol levels were associated with lower hippocampal PVS burden (95% CI=0.803–0.955, *p*_FDR_=0.028). Within the genetic domain, several PRS showed region-specific associations. In the midbrain, higher genetic risk factor for depressive disorder was associated with increased PVS counts (95% CI=1.047–1.285, *p*_FDR_=0.019), while higher genetic risk factor for insomnia was associated with lower PVS counts (95% CI=0.797–0.972, *p*_FDR_=0.022). In the hippocampus, higher polygenic risk for anxiety and stress-related disorders were associated with lower PVS counts (95% CI=0.824–0.985, *p*_FDR_=0.044 for both) [**Supplementary Table 9**].

In individuals with abnormal amyloid but normal tau (A+T-), demographic determinants were significantly associated with regional PVS counts, while vascular determinants exhibited a nominal significance. Sex remained a strong predictor, with female sex associated with higher hippocampal PVS counts (95% CI=1.126–2.016, *p*_FDR_=0.032) [**Supplementary Table 10**].

In participants with both amyloid and tau pathology (A+T+), distinct associations emerged. Sex remained a significant predictor, with female sex associated with lower centrum semiovale-PVS counts (95% CI=0.193–0.703, *p*_FDR_=0.013). Strong effects were also observed for former smoking, associated with higher centrum semiovale-PVS counts (95% CI=2.378–15.910, *p*_FDR_=0.003) and sleep duration, associated with higher hippocampal counts (95% CI=1.079–1.762, *p*_FDR_=0.05) [**Supplementary Table 11**].

### 3.6 Sensitivity analyses

When additionally adjusting for the volume of the corresponding anatomical region, the results were largely consistent with the primary analyses. In some cases, the magnitude of the associations became stronger after volume adjustment, including the higher PVS burden in females in the hippocampus, midbrain and basal ganglia, and the positive association between age and centrum semiovale PVS. Others were modestly attenuated but remained statistically significant [**Supplementary Tables 12-22**].

## 4. Discussion

In this large cohort of CU individuals at increased genetic risk for AD, we identified multidomain determinants of PVS burden across distinct brain regions. Demographic, cardiovascular, lifestyle/environmental and genetic factors contributed differentially to regional PVS variation, with several associations moderated by sex, amyloid status and AD pathology stage. These findings highlight the complexity of PVS determinants and support their potential as early MRI markers of cerebrovascular and glymphatic vulnerability in preclinical AD.

The centrum semiovale emerged as the most consistently affected region across demographic and genetic domains, reinforcing prior evidence that MRI-visible PVS in this region are closely linked to the accumulation of Aβ pathology, which is a hallmark of the AD continuum [8,10,11,47]. In contrast, the basal ganglia showed a different etiological profile, showing a strong association with BMI, which is maintained in the men-stratified analysis, consistent with its sensitivity to hypertensive arteriopathy [8,11,47]. The hippocampal and midbrain PVS burden showed distinct patterns, with associations more tightly linked to AD biomarkers and psychiatric or sleep-related factors, suggesting that perivascular dysfunction in these regions may be driven by early glymphatic and neuroinflammatory processes rather than traditional vascular mechanisms [1,3,48]. The hippocampus, in particular, is known to be highly sensitive to sleep-related disruption of perivascular drainage [49]. This spatial pattern supports the notion that distinct perivascular mechanisms operate across vascular and limbic territories, reflecting the interplay between hemodynamic stress, metabolic demand, and clearance pathways [7,8,10,11,47]. Together, these results point to regional specificity in the mechanisms underlying PVS counts, vascular and hemodynamic influences in white-matter regions such as the centrum semiovale and basal ganglia, and clearance or inflammatory dysfunction in limbic regions such as the hippocampus and midbrain.

Age was a robust determinant of PVS counts, but its effects were region-specific. Advancing age was significantly associated with greater PVS burden only in the centrum semiovale, whereas no significant effects were found in the basal ganglia, hippocampus, or midbrain. This finding differs from large population-based studies, which reported age-related PVS increases across all regions, particularly in the basal ganglia [7,17]. The narrower age range and younger mean age of the ALFA cohort could likely explain this regional restriction, as white-matter PVS tend to appear earlier in life, while basal ganglia PVS emerge later with cumulative vascular injury [50]. Our results therefore suggest that centrum semiovale perivascular alterations may represent early manifestations of vascular stress in midlife, potentially preceding the deeper basal changes seen in older or clinically affected populations.

Sex also emerged as a consistent determinant of PVS burden. Men exhibited higher PVS counts in the centrum semiovale, whereas women showed higher counts in the other three regions. This pattern aligns partially with previous large-scale imaging studies that found higher centrum semiovale and midbrain PVS counts in men [7,51], higher hippocampal PVS counts in women [7,52] and varying results for basal ganglia PVS counts [7,52,53,54]. These differences may reflect earlier, sex and AD-specific influences of vascular and hormonal factors in our younger, healthier sample that are not yet detectable in general population samples. Notably, the predominance in hippocampal and midbrain burden in women may reflect greater vulnerability of estrogen-responsive regions, as post-menopauseal declines in estradiol are known to impair neuroinflammatory regulation, mitochondrial function, sleep architecture and glymphatic clearance [55] while the stronger centrum semiovale effects in men may correspond to higher vascular stiffness and blood pressure profiles. Notably, the presence of more PVS in the hippocampus of women is in line with their higher lifetime risk of AD and hypotheses that suggest greater depositions of Aβ in the hippocampus and cortical tissues, as pointed by others [7].

Sex-stratified analyses further supported the existence of sex-specific biological pathways influencing PVS burden. The most robust interaction was observed between sex and the PRS for HDL cholesterol in the midbrain, indicating that genetic variation in lipid metabolism affects perivascular integrity differently in men and women. In men, a higher PRS for HDL cholesterol was associated with greater midbrain PVS counts, whereas in women, a higher PRS for HDL cholesterol predicted lower centrum semiovale PVS burden. These results point towards distinct regulation of lipid transport and cerebrovascular homeostasis, consistent with prior evidence linking estrogen, lipid metabolism and vascular function [56,57].

Cardiovascular risk factors were key contributors to PVS burden, supporting the close relationship between vascular health and perivascular function [7,17,53]. Blood pressure emerged as the main cardiovascular risk factor. Both SBP and, in particular, DBP were positively associated with PVS counts in the centrum semiovale and midbrain. This finding is consistent with previous evidence from the general population, which suggests that DBP is a particularly sensitive determinant of PVS and may directly influence perivascular fluid exchange more than SBP [7]. These effects persisted after adjustment for age and sex, and remained consistent after stratification by AD biomarkers, indicating that vascular factors contribute to perivascular enlargement independently of amyloid or tau pathology. Interestingly, while previous studies reported that cardiovascular risk factors were most strongly associated with hippocampal PVS [7], we did not observe significant associations in the main-analysis in this region. However, in amyloid-specific models, we showed a nominal interaction between SBP and amyloid status in the hippocampus, with higher SBP associated with greater hippocampal PVS burden in Aβ- individuals. The overall lower blood pressure levels in ALFA, could be partly accounting for these weaker and regionally restricted associations. In addition, our findings are consistent with the hypothesis that vascular dysfunction and impaired arterial pulsatility hinder glymphatic clearance, leading to solute accumulation and PVS dysfunction [12,58].

Lifestyle and mental health factors further modulated PVS burden with region-specific patterns. We observed a significant interaction between depression and Aβ status for hippocampal PVS, suggesting that depressive symptoms confer additional burden in an amyloid!zldependent manner. Greater physical activity was associated with reduced centrum semiovale PVS burden, and this association persisted in Aβ+ individuals. In addition, longer sleep duration was related to higher hippocampal PVS burden in biomarker-positive participants. These observations were consistent with literature linking disturbed sleep with impaired glymphatic clearance [49,59]. In the main analysis, an insomnia PRS was associated with lower hippocampal PVS, whereas no consistent genetic effects for sleep-related traits were observed, suggesting that sleep’s influence on perivascular function is primarily behavioral or environmentally mediated rather than genetically determined in a cohort of middle aged at risk individuals for AD. Smoking and air pollution also contributed to PVS burden. Former smoking was associated with higher centrum semiovale burden in A+T+ participants, while PM_2.5_ exposure showed AT-dependent associations, reinforcing the notion that chronic systemic inflammation and vascular stress exacerbate perivascular dysfunction [1,60,61,62].

Genetic factors also played an important and independent role in shaping PVS burden. PRS for metabolic traits, especially BMI and HDL cholesterol, were linked to lower PVS counts in the centrum semiovale. These associations contrast with expectations based on vascular risk but are consistent with the complex and sometimes paradoxical relationships reported between BMI and cerebrovascular or neurodegenerative outcomes [63,64]. In support of these heterogeneity, previous work [7] also reported opposite effects of BMI across regions (higher hippocampal PVS with manual counts but lower with total counts), a pattern that could be reflecting demographic differences in the manually counted subset, in addition to anatomical and methodological factors.

Distinct genetic signatures were also evident for some biomarker stages. In participants without amyloid or tau pathology, psychiatric and sleep-related PRS (anxiety, depression, insomnia) showed associations with hippocampal and midbrain burden, whereas in Aβ+ individuals, *APOE-*ε4 became more prominent in the hippocampus, with higher PVS counts associated with both ε4 carriership and single-allele load, corroborating general population-based findings, where dose-dependent associations between *APOE*-ε4 allele count and hippocampal PVS burden were detected [7]. In those within the A+T+ group, no PRS signal survived FDR correction. These shifts suggested that different genetic mechanisms predominate along the AD *continuum*, from psychological pathways in the earliest stages [65,66,67], to *APOE*-related influences once amyloid is present, although larger stratified datasets will be needed to draw firmer conclusions [68,69].

The observed findings have several implications for early detection and prevention. PVS burden, which is now measurable on standard MRI via automated methods, may serve as a low-cost biomarker capturing the cumulative impact of vascular, lifestyle, and genetic risk in preclinical AD. The identification of modifiable factors highlights concrete prevention targets. Furthermore, the differential effects by sex and biomarker stage suggest that strategies to preserve cerebrovascular health should be tailored to individual risk profiles, integrating biological sex and AD biomarker status [18,70]. This aligns with emerging precision medicine approaches aimed at delaying or preventing symptomatic conversion.

This study has several strengths. It leverages the well-characterized ALFA cohort, a deeply phenotyped, biomarker-enriched sample of CU adults in midlife, providing a unique opportunity to investigate early cerebrovascular and glymphatic alterations. The use of a fully automated, reproducible quantification pipeline for PVS ensures objective and region-specific assessments, improving sensitivity compared to visual scales. Integrating multidomain risk profiling allowed us to disentangle independent and interacting determinants of PVS burden in unprecedented detail. Importantly, additional sensitivity analyses adjusting for the volume of the corresponding anatomical regions yielded highly consistent results, supporting the robustness of the primary findings and indicating that the observed associations were not driven by differences in regional brain volume. The large sample size and harmonized imaging and biomarker protocols enhance internal validity and comparability with other AD research preclinical initiatives. Nonetheless, several limitations should be acknowledged. The cross-sectional design precludes causal inference and limits interpretation of temporal relationships between PVS burden, making it impossible to determine whether PVS burden precedes or follows early AD-related changes. While the automated quantification reduces subjectivity, it remains sensitive to scanner-specific variability and technical artifacts, warranting replication across diverse populations and imaging platforms. Although we corrected for multiple testing, some associations remained at nominal significance and should be interpreted as exploratory. Furthermore, the genetic analyses are constrained by current PRS methods, which do not fully capture rare variants or gene-environment interactions that may influence PVS. In addition, stratified genetic analyses, particularly those involving *APOE*!zlε4 homozygotes, were limited by small subgroup sample sizes, which may have reduced power to detect associations and contributed to marginal or non-significant effects. Finally, the ALFA cohort is genetically and demographically enriched for AD risk and relatively homogeneous in terms of ancestry, which may limit the generalizability of our findings to the general population.

Future work should examine longitudinal trajectories of PVS burden in relation to AD biomarker progression, cognitive performance, and clinical outcomes. Integrating PVS with other markers of CSVD, such as white matter hyperintensities, could help disentangle shared and distinct vascular pathways contributing to impaired brain clearance. Combining MRI-based PVS measures with complementary glymphatic proxies, including diffusion tensor-imaging-based analysis along the perivascular space (DT-ALPS) [71], and fluid biomarkers of neuroinflammation, vascular injury, and metabolic stress could clarify mechanistic pathways linking vascular and glymphatic dysfunction to AD risk. Intervention studies targeting vascular and lifestyle risk factors are essential to determine whether interventions on those risk factors can modify PVS trajectories and reduce subsequent cognitive decline. Ultimately, integrating perivascular imaging metrics into multimodal biomarker frameworks may refine risk stratification and accelerate the development of personalized prevention strategies in AD.

## 5. Conclusions

PVS are MRI-visible markers of glymphatic and vascular clearance function. In this cohort of CU individuals with a genetic predisposition for AD, PVS burden was influenced by demographic, vascular, lifestyle, and genetic factors, with distinct patterns by sex and AD biomarker status. Our findings revealed that multidomain risk exposures and inherited metabolic vulnerability jointly modulate early cerebrovascular dysfunction before symptom onset. Because PVS quantification is feasible in standard MRI, it represents a promising tool for individualized risk stratification and precision prevention in preclinical AD.

## Data Availability

De-identified data supporting the findings of this study are available upon request from the corresponding author on reasonable request (NV-T). Requests are evaluated by the Scientific Committee at the Barcelonaβeta Brain Research Center and, if granted, data are shared and regulated by a Data Sharing Agreement.

## 6. List of abbreviations

AIC: Akaike Information Criterion
ALFA: Alzheimer’s and Families
AD: Alzheimer’s Disease
AT: Amyloid/Tau
Aβ: Amyloid-β
*APOE*: Apolipoprotein E
BFI: Body Fat Index
BMI: Body Mass Index
CAIDE: Cardiovascular risk factors, Aging and Incidence of Dementia
CSF: Cerebrospinal Fluid
CU: Cognitively Unimpaired
CI: Confidence Interval
DBP: Diastolic Blood Pressure
FDR: False Discovery Rate
HRC: Haplotype Reference Consortium
HDL: High-Density Lipoprotein cholesterol
LDL: Low-Density Lipoprotein cholesterol
MRI: Magnetic Resonance Imaging
MET: Metabolic Equivalent of Task
PM: Particulate Matter
PVS: Perivascular Spaces
PRS: Polygenic Risk Score
RR: Rate Ratio
SBP: Systolic Blood Pressure

## 7. Declarations

## Ethics approval and consent to participate

The ALFA study protocol was approved by the Independent Ethics Committee Parc de Salut Mar Barcelona and registered at Clinicaltrials.gov (Identifier: NCT01835717). It was conducted in accordance with the directives of the Spanish Law 14/2007, of 3rd of July, on Biomedical Research (Ley 14/2007 de Investigación Biomédica). ALFA study participants accepted the study procedures by signing an informed consent form and had a close relative volunteering to participate in the functional assessment procedure of the participant, who also granted his or her consent. A prerequisite for entering the ALFA+ study (Clinicaltrials.gov Identifier NCT02485730) was accepting the study procedures and understanding that the CSF results will not be disclosed.

## Consent for publication

Not applicable.

## Competing interests

The authors declare no competing interests.

## Funding

GS-B is supported by the Instituto de Salud Carlos III (ISCIII) through the project CP23/00039 (Miguel Servet contract), co-funded by the European Union (FSE+) and by the Spanish Research Agency through the project PID2024-163071OB-I00, funded by MICIU/AEI/10.13039/501100011033 and by the European Union FEDER.

TEE is supported by the Alzheimer Association Research postdoctoral fellowship 25AARF-1377279, Alzheimer Nederland project WE.03-2024-07, and Williams H. Gates Sr. AD Fellowship from the Alzheimer’s Disease Data Initiative. This project is partly supported by the National Computing Facilities Processing research programme which is financed by the Dutch Research Council (NWO) under the grant [2022.018].

NV-T acknowledges support from the Ramón y Cajal Fellowship (RYC2022-038136-I) and the project PID2022-143106OA-I00, both funded by MCIN/AEI/10.13039/501100011033, with co-funding from FSE+ and FEDER, EU respectively. Additionally, NV-T received support from the Williams H. Gates Sr. AD Fellowship from the Alzheimer’s Disease Data Initiative, and the Alzheimer’s Association, AD Strategic Fund (Project #VCID-UMD-26-1514428).

## Author Contributions

AF-B: data analysis, writing (original draft), interpretation of the data. GT-S: writing (original draft), interpretation of the data. PG: writing (review and editing). BR-F: writing (review and editing). ED-G: writing (review and editing). JH: data acquisition curation, neuroimaging. MTB: writing (review and editing). GSB: writing (review and editing). MC: data acquisition and curation of air pollution data, writing (review and editing). MN: writing (review and editing). MdB: Method development, writing (review and editing). TEE: conceptualization, neuroimaging data acquisition and curation, writing (review and editing). NV-T: conceptualization and design of the study, supervision, writing (original draft), interpretation of the data. All authors had full access to all the data in the study, reviewed and edited the manuscript, approved the final version of the manuscript, and had final responsibility for the decision to submit for publication.

## Acknowledgements

This publication is part of the ALFA study (ALzheimer and FAmilies). The authors would like to express their most sincere gratitude to the ALFA project participants and relatives without whom this research would not have been possible.

## Notes

### Competing Interest Statement

The authors have declared no competing interest.

### Clinical Trial

NCT01835717

### Author Declarations

The ALFA study protocol was approved by the Independent Ethics Committee Parc de Salut Mar Barcelona and registered at Clinicaltrials.gov (Identifier: NCT01835717).

