## Supplementary figures and images for "Multidomain determinants of perivascular spaces indicate early cerebrovascular vulnerability in individuals at risk of Alzheimer’s disease"

### Supplementary Figure 1

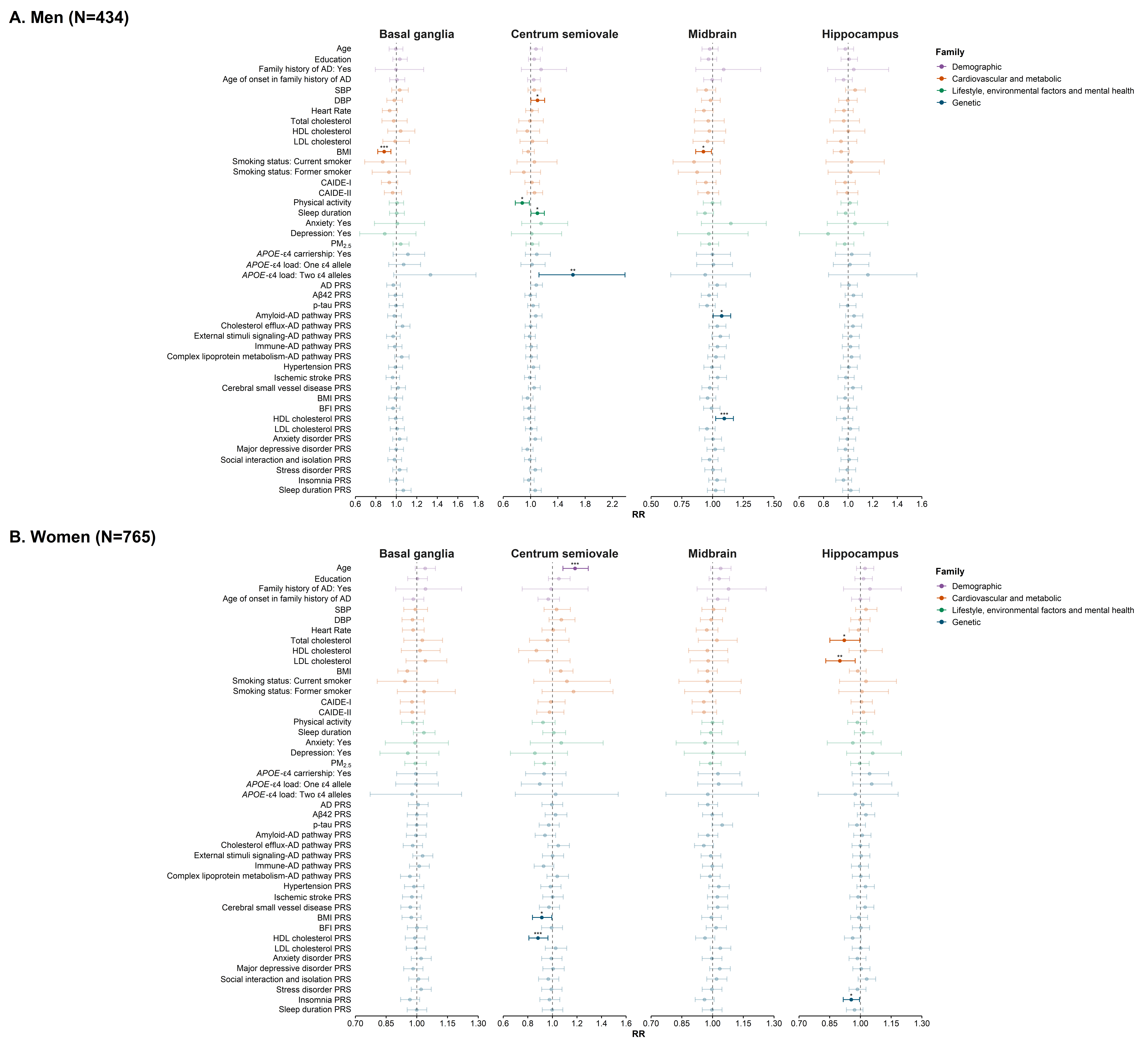

### Supplementary Figure 2

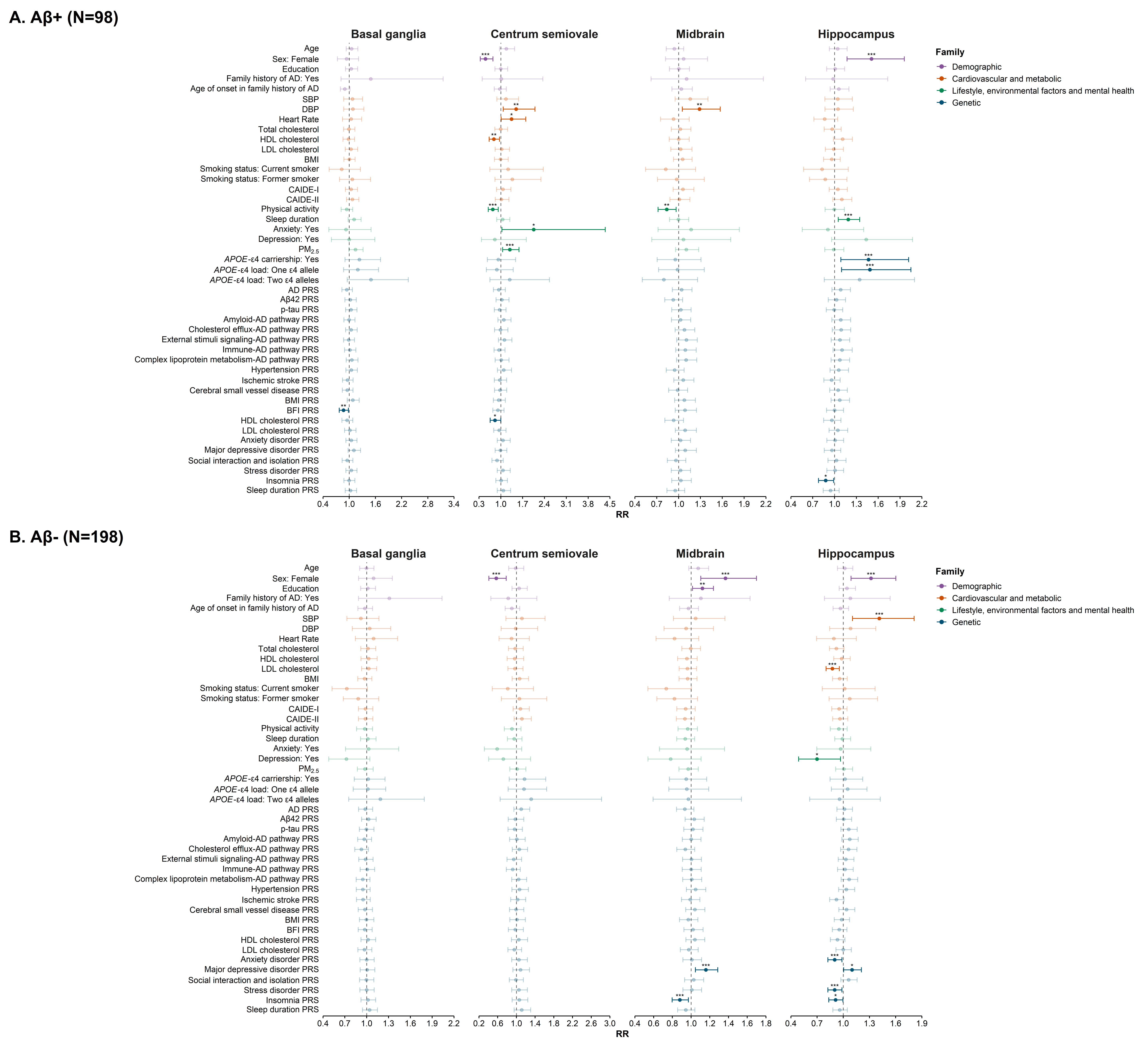

### Supplementary Figure 3

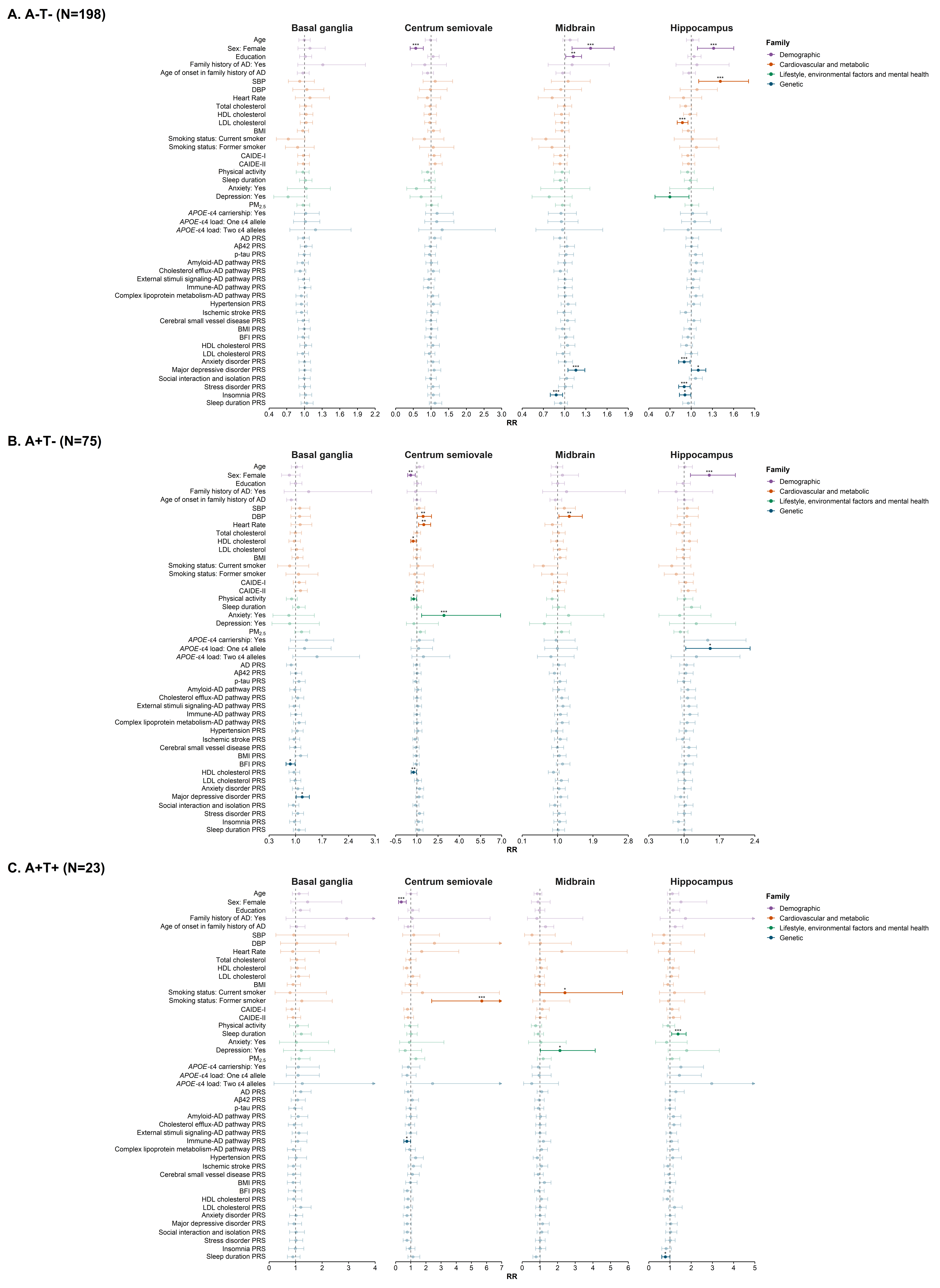
